# Intraocular pressure, primary open-angle glaucoma and the risk of retinal vein occlusion: a Mendelian randomization mediation analysis

**DOI:** 10.1101/2024.01.31.24302094

**Authors:** Andreas Katsimpris, Sebastian-Edgar Baumeister, Nafsika Voulgari, Hansjörg Baurecht, Stylianos Kandarakis, Michael Nolde

## Abstract

**Background:** The etiological connection between intraocular pressure (IOP) and the risk of retinal vein occlusion (RVO) remains elusive, particularly regarding whether this risk emanates from the direct influence of elevated intraocular pressure (IOP), irrespective of the presence of primary open-angle glaucoma (POAG), or if it arises as a consequence of the sequelae of POAG. Therefore, we conducted a Mendelian Randomization (MR) mediation analysis to elucidate the mediating role of POAG in the association between IOP and RVO.

**Methods:** We identified 47 single-nucleotide polymorphisms (SNPs) associated with IOP (P-value < 5×10^-8^) leveraging data from a genome-wide association study (GWAS) (N = 97,653) obtained from the UK Biobank and 50 SNPs associated with POAG (P-value < 5×10^-8^) from a GWAS meta-analysis (16,677 cases and 199,580 controls). We related these SNPs with RVO using a GWAS of 775 RVO cases and 376,502 controls from FinnGen. By utilizing univariable and multivariable MR analyses we calculated the total effect of IOP on RVO and estimated the degree to which POAG mediates this association.

**Results:** MR analyses showed that higher IOP is associated with higher RVO risk (odds ratio of RVO per 1 mmHg increase in IOP: 1.53; 95% confidence interval: 1.04 to 2.26; p-value = 0.03). Moreover, our MR mediation analysis suggested that 91.6% of the total effect of IOP on RVO risk was mediated through POAG. The primary results were consistent with estimates of pleiotropy-robust MR methods.

**Conclusion:** Our findings suggest that higher IOP increases the risk of RVO and that the majority of this effect is mediated through POAG.

## Introduction

Retinal vein occlusion (RVO) is the second most common retinal vascular disease following diabetic retinopathy and an important cause of vision loss in individuals older than 80 years with a prevalence of 4.6% [1]. Risk factors for RVO are mainly related to atherosclerosis, but also conditions altering the rheologic properties in the retinal veins, such as hypercoagulability and vasculitis [2].

Glaucoma, an ocular disorder characterized by optic nerve damage and visual field loss, has been implicated in the pathogenesis of RVO [3]. Specifically, the association between RVO and primary open angle glaucoma (POAG) has been investigated in numerous studies, showing a significantly higher prevalence of RVO in patients with POAG compared to the general population [3,4]. Different pathogenic mechanisms have been proposed, implicating intraocular pressure which is a well-established and modifiable risk factor for glaucoma. On one hand, increased IOP has been hypothesized to collapse the retina vessel walls thus resulting in occlusion [5,6]. On the other hand, structural changes of the vessels at the glaucomatous optic discs might also make them more susceptible to thrombosis [7–10]. Whether RVO is caused by the effects of increased IOP independently from any glaucomatous optic disc abnormalities has not been determined.

Mendelian randomization (MR) serves as a robust methodology utilizing genetic variants derived from genome-wide association studies (GWAS) as instrumental variables to elucidate the causal relationship between modifiable exposures and disease outcomes [11]. A recent MR study has identified a positive association between POAG and RVO [12]. However, it was not assessed whether this observed effect is intrinsic to POAG itself or if it is a consequence of the elevated intraocular pressure that is mediated through POAG. In addressing this gap, we conducted a MR mediation analysis to discern the mediating role of POAG in the relationship between IOP and RVO and sought to enhance our understanding of the nuanced interplay between these factors.

## Materials and Methods

### Study design

MR employs genetic variants as instrumental variables to investigate the impact of risk factors on disease susceptibility [11]. This method mitigates susceptibility to confounding and reverse causation, as these genetic variants are randomly assigned at conception, creating a quasi-randomized exposure allocation analogous to a randomized trial [11]. In our investigation, a two-sample MR was executed using summary statistics of single-nucleotide polymorphisms (SNPs) obtained from GWAS focusing on IOP [13] and RVO [14]. The aim was to evaluate the influence of IOP on the risk of RVO. Moreover, we tried to assess the proportion of the effect of IOP on RVO that is potentially mediated through POAG, by employing a two-step MR for mediation analysis [15]. The study protocol was not pre-registered and our research adhered to the STROBE-MR guidelines [16] and “Guidelines for performing Mendelian randomization investigations” [17].

### Data sources

Summary data for corneal-compensated IOP were obtained from the UK Biobank (UKBB) cohort GWAS [13] (Supplementary Table 1). In particular, a subset comprising 97,653 individuals within the UKBB underwent ophthalmic evaluations, encompassing the quantification of corneal-compensated IOP measured in millimeters of mercury (mmHg) through the application of a non-contact tonometer. The preference for corneal-compensated IOP as the selected exposure phenotype over regular IOP stems from its designed capability to accommodate corneal biomechanical properties. Moreover, this metric has been previously employed in GWAS for IOP [18]. Summary data for RVO were obtained from the FinnGen consortium database’s R9 release, involving a cohort of 775 documented RVO cases and 376,502 controls [14]. The FinnGen GWAS participants were of European origin, and the identification of RVO cases adhered to diagnostic criteria established in either the International Classification of Diseases, Ninth Revision (ICD-9) or International Statistical Classification of Diseases, Tenth Revision (ICD-10) code. For the mediation MR analysis, GWAS summary statistics for POAG were also retrieved from a GWAS meta-analysis of 16,677 POAG cases and 199,580 controls of European ancestry [19] (Supplementary Table 1). POAG cases were selected based on the ICD-9/ICD-10 criteria [19]. The genotyping, quality control, and imputation methods applied to the GWAS data used in our study have been detailed elsewhere [14,19,20].

### Selection of genetic variants as instrumental variables

We selected SNPs identified in the IOP GWAS that achieved genome-wide significance (P-value < 5*10^-8^), following clumping for linkage disequilibrium (LD) at r^2^ < 0.001 over a 10mb window [17]. The MR-Steiger directionality test was employed to discern the directions of the causal relationship between IOP and RVO [21]. SNPs exhibiting a stronger correlation with the outcome than the exposure were systematically excluded, as were those demonstrating notable influence in the funnel plots and scatter plots. Ultimately, we identified 47 SNPs associated with IOP as instrumental variables. Furthermore, we quantified the proportion of variability in IOP that is explained by these 47 SNPs, through the summation of the coefficients of determination (R^2^) derived from the associations between the selected SNPs and IOP. Employing a similar approach, we chose 50 SNPs from the GWAS on POAG for utilization in our MR mediation analysis.

### Statistical analysis

After the process of data harmonization based on HapMap3 [22], the removal of strand-ambiguous variants, and the alignment of association estimates, we proceeded to calculate Wald ratios. We calculated these ratios by dividing the logarithm of odds ratio per allele for each SNP identified in the GWAS for RVO by its respective estimate obtained from the IOP GWAS. Subsequently, the total effect of IOP on RVO risk was assessed through a multiplicative random effects inverse variance weighted (IVW) meta-analysis of the Wald ratios [23].

We implemented a univariable two-sample MR methodology, utilizing summary data extracted from GWAS of IOP and RVO. Our two-sample MR analysis was based on three key assumptions. Firstly, the imperative that the genetic variants selected should be associated with the targeted risk factor, a principle known as the “relevance” assumption [24]. We adhered to this assumption by choosing SNPs as instrumental variables that reached the threshold of genome-wide significance (P-value < 5*10^-8^). Furthermore, we evaluated the strength of our instrumental variables by assessing the F-statistic of the selected SNPs, concurrently examining the proportion of variance in the exposure that they accounted for [25]. Secondly, the selected genetic instruments should not be correlated with factors that could potentially confound the relationship between the exposure and outcome, also known as the “exchangeability” assumption [24]. Thirdly, the “exclusion restriction” assumption [24], where the genetic instruments should not influence the outcome except through the risk factor of interest. While the “exchangeability” and “exclusion restriction” assumptions are inherently unverifiable, we executed sensitivity analyses to identify potential violations of these MR assumptions.

PhenoScanner [26] was employed to assess whether any of our selected genetic instruments were associated with phenotypes that could serve as potential confounders in our analysis. In instances where pleiotropic pathways were identified, we applied multivariable MR to adjust for these effects [27]. Moreover, we examined heterogeneity among our selected SNPs using the Cochran Q heterogeneity test and IGX^2^ [28] to detect pleiotropy, utilized several MR methods proposed to enhance robustness in instances where genetic variants exhibit pleiotropy [29] (MR Egger regression, penalized weighted median, IVW radial regression, and MR-Pleiotropy Residual Sum and Outlier (MR-PRESSO)) and performed a leave-one-out analysis to ascertain if the IVW estimate was influenced by a singular SNP.

To further explore the mediating effect of IOP on RVO through POAG, we conducted a two-step MR for mediation analysis, as outlined in previous literature [15]. This process involves the calculation of two distinct MR estimates: firstly, calculating the effect of IOP on POAG through a univariable MR model, and secondly, assessing the effect of POAG on RVO using a multivariable MR model that includes an adjustment for IOP. The IOP estimate on POAG multiplied with the adjusted POAG estimate on RVO provided the indirect effect of IOP on RVO, mediated through POAG. We also quantified the proportion of the total effect of IOP on RVO explained by the mediator (POAG) by dividing the calculated indirect effect of IOP on RVO by the total effect (adjusted direct and indirect effects of IOP on RVO). The delta method was employed to derive 95% confidence intervals (95%CI) for the indirect effect [30]. Furthermore, considering the inherent requirement for independence between the SNPs selected as instruments for the exposure (IOP) and mediator (POAG) in MR mediation analysis, we ensured that the selected SNPs from the IOP and POAG GWAS datasets were non-overlapping [15].

This precautionary step was essential to uphold the validity and integrity of the MR for mediation analysis, preventing potential bias arising from SNP overlap.

We conducted all analyses with R version 4.2.1 [31] using the MVMR (0.3), TwoSampleMR (0.5.6), MendelianRandomization (0.5.1), MRPRESSO (1.0) and cause (1.2.0) packages.

## Results

The 47 SNPs selected from the IOP GWAS (Supplementary Figure 1) accounted for 2.64% of the variability in IOP, with all SNPs displaying F-statistics of ≥29.96 (Supplementary Table 2). Employing the IVW method, genetically predicted IOP was positively associated with RVO risk (OR = 1.53 per 1mmHg increase in IOP; 95%CI = 1.04 to 2.26; P-value = 0.03) (Figure 2 and Supplementary Figure 2). Results from pleiotropy-robust MR methods aligned with the IVW analysis estimate (Figure 2). We found no associations of our selected SNPs with RVO risk factors apart from POAG (Supplementary Table 3) and, thus, we refrained from conducting multivariable MR to correct for potential correlated horizontal pleiotropy.

**Figure 1.**
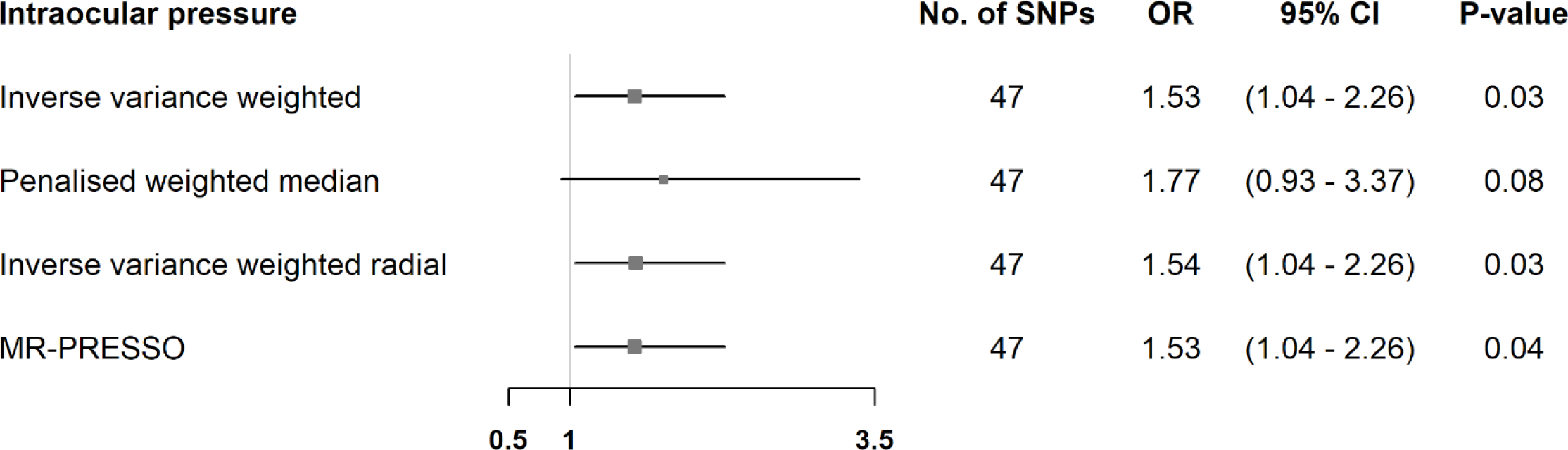
Mendelian randomization estimates for the effect of intraocular pressure on retinal vein occlusion. Estimates are reported as changes in odds of retinal vein occlusion per 1 mmHg increase in intraocular pressure. SNP, single nucleotide polymorphism; CI, confidence interval; MR-PRESSO, Mendelian randomization pleiotropy residual sum and outlier.

**Figure 2.**
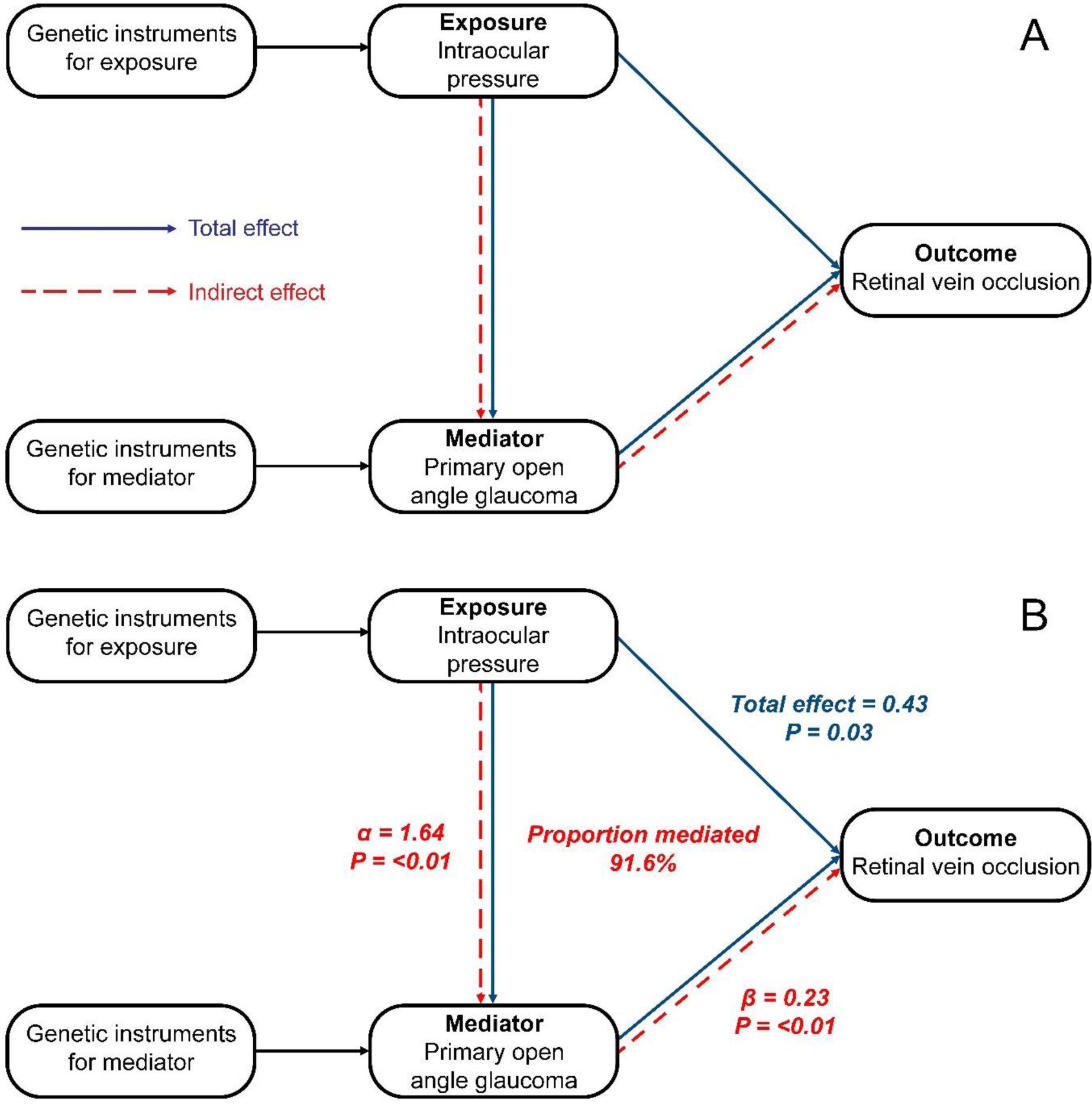
Directed acyclic graphs of the mediation analysis with Mendelian randomization. The indirect effect of intraocular pressure (IOP) on retinal vein occlusion (RVO) can be calculated by multiplying α times β, where α is the effect of IOP on primary open-anlge glaucoma (POAG), and β the effect of POAG on RVO. The proportion mediated can be estimated by dividing the indirect effect by the total effect of IOP on RVO. All estimates are shown as the difference in the logarithm of odds of the outcome, per 1 unit increase of the exposure (continuous variables: IOP) or between the two exposure groups (binary variables: POAG).

The Wald ratios for IOP with RVO did not exhibit heterogeneity (Supplementary Table 4) and the Cochran’s Q heterogeneity test yielded a value of 37.89 (p-value = 0.797). Additionally, no evidence for directional pleiotropy was found, since the intercepts from the MR-Egger analyses did not deviate from zero (Supplementary Table 4). The leave-one-SNP-out analysis did not identify any SNPs exerting significant influence on the IVW estimate for the association between IOP and RVO risk (Supplementary Table 5).

An illustrative depiction of the MR mediation analysis can be seen in Figure 2. We found that 91.6% of the total effect of IOP on RVO was mediated through POAG (Supplementary Table 6).

## Discussion

In this MR mediation analysis, we utilized genetic data to disentangle the causal pathway between IOP and RVO, and to also explore the mediating role of POAG in this relationship. Our results support a positive causal effect of IOP on RVO which is mostly mediated through POAG.

Many studies have examined the relationship between glaucoma, specifically POAG, and risk for RVO [4]. Yin et al demonstrated in their meta-analysis that POAG is significantly associated with RVO risk (OR: 5.03; 95% CI: 3.97 to 6.37) [3]. Moreover, in their subgroup analysis, POAG and chronic open angle-glaucoma were also correlated with central RVO (OR: 13.30; 95% CI: 3.34 to 53.20) and branch RVO (OR: 2.14; 95% CI: 1.09 to 4.20), but conclusions are guarded due to the small number of studies included in the meta-analyses. Similarly, Na et al. determined that the RVO incidence rate for the open-angle glaucoma patients was 528.95 per 100,000 person-years (95% CI: 515.46 to 542.79), 3.27 times higher than in the general population [32]. Another Korean nationwide, population-based 11-year longitudinal study showed that patients with POAG were under higher risk for developing RVO (HR: 3.27, 95% CI: 2.55-4.19) [33], while in the Beaver Dam Eye Study, no significant difference was found in the age-adjusted 5-year incidence of RVO between participants with and without POAG [34].

Different potential mechanisms have been proposed to interpret the association between POAG and RVO, and IOP has been shown to be a major contributing factor affecting retinal hemodynamics. It has been suggested that increased IOP may compress blood vessels, leading to proliferation of the vein intima and subsequently to the collapse of retinal vessel walls [5]. The effects of this mechanical compression are more pronounced at the level of the lamina cribrosa where the venous pressure is lowest. In older individuals with impaired retinal circulation and compromised autoregulatory vascular mechanisms, even a slight increase in IOP could be significant enough to decrease ocular blood flow, leading to venous slowing or stasis [6]. Luntz et al. strongly support an etiological relationship between increased IOP and central RVO having shown that the incidence of RVO in ocular hypertension is similar to that of POAG [35]. However, the Ocular Hypertension Treatment study, which included only participants with ocular hypertension, did not find a significant difference in RVO incidence between the observation and medication group [36].

The effect of IOP on retinal vessels might be further accentuated by anatomical characteristics of the glaucomatous optic nerve head. The posterior bowing of the lamina cribrosa present in glaucomatous optic discs confers less glial support to the retinal vessels thus making them more vulnerable to changes in IOP [8]. Also, higher cup-to-disc ratio has been correlated with the development of RVO, postulating that optic disc cupping induces structural abnormalities to the vessel at the disc, thus altogether making them susceptible to occlusion, venous stasis, and, consequently, to thrombosis, according to Virchow’s triad [7,9,10]. On top of that, according to the vascular theory of glaucoma pathogenesis, patients with POAG have narrower retinal arteries and veins compared to normal individuals, predisposing them to greater vessel wall compression following a rise in IOP [37]. Moreover, it has been hypothesized that RVO and POAG could potentially arise from the same vascular abnormality given their shared vascular risk factors. Optic disc hemorrhages frequently encountered in POAG and RVO indicate small vein occlusions, raising the question of a common underlying pathway [38].

The key strength of this study is that it stands out as the first study to utilize the approach of MR mediation analysis to disentangle the complex causal pathway between IOP, POAG and RVO. Another strength of it lies in the consistency of association estimates obtained from pleiotropy-robust methods, aligning with the IVW estimate and indicating an absence of model violations. Nevertheless, certain limitations warrant consideration. Firstly, our investigation primarily focused on delineating the mediating role of POAG in the association between IOP and RVO, excluding exploration into other glaucoma subtypes such as primary angle-closure glaucoma.

Secondly, our MR models assumed a linear relationship between the identified risk factors and the observed outcomes, whereas the true association among IOP, POAG, and RVO may manifest as non-linear. Thirdly, it is imperative to exercise caution when extrapolating the genetic associations derived from European populations to other ethnic groups due to potential population-specific variations. Fourthly, we did not assess the associations of IOP with the two main subtypes of RVO, namely branch RVO and central RVO, since no GWAS datasets are available for these phenotypes.

In conclusion, our MR analyses infer an elevated risk of RVO associated with higher IOP. By utilizing MR mediation analysis, our findings additionally suggest that the majority of the effect of IOP on RVO risk is mediated through POAG. To advance our understanding of this intricate interplay, further investigations through population-based prospective studies, as well as experimental studies, are warranted to comprehensively explore the complex pathway involving IOP, POAG, and RVO.

## Author contributions

KA, BSE and NM contributed to the study conception and design, drafted the manuscript and analyzed the data. All authors critically revised the manuscript for important intellectual content, provided administrative, technical, or material support and approved the final version.

## Data availability

The summary statistics for the intraocular for the UKBB GWAS are available at https://pan.ukbb.broadinstitute.org (access date: 2023/10/12). The retinal vein occlusion summary statistics for the FinnGen GWAS are available at https://www.finngen.fi/en/access_results (access date: 2023/10/12). The primary open-angle glaucoma summary statistics are available at https://www.ebi.ac.uk/gwas/publications/33627673 (access date: 2023/10/12)

## Additional information

### Competing Interests

None declared

## Data Availability

All data produced in the present work are contained in the manuscript

## Acknowledgments

We want to acknowledge the participants and investigators of the FinnGen and UK Biobank studies.

## Notes

### Competing Interest Statement

The authors have declared no competing interest.

### Funding Statement

This study did not receive any funding

