## Supplementary tables and figures for "Intraocular pressure, primary open-angle glaucoma and the risk of retinal vein occlusion: a Mendelian randomization mediation analysis"

Supplementary Figures and Tables

**Supplementary Figure 1** Funnel plot of single SNP Wald ratio estimates for the effect of intraocular pressure on retinal vein occlusion

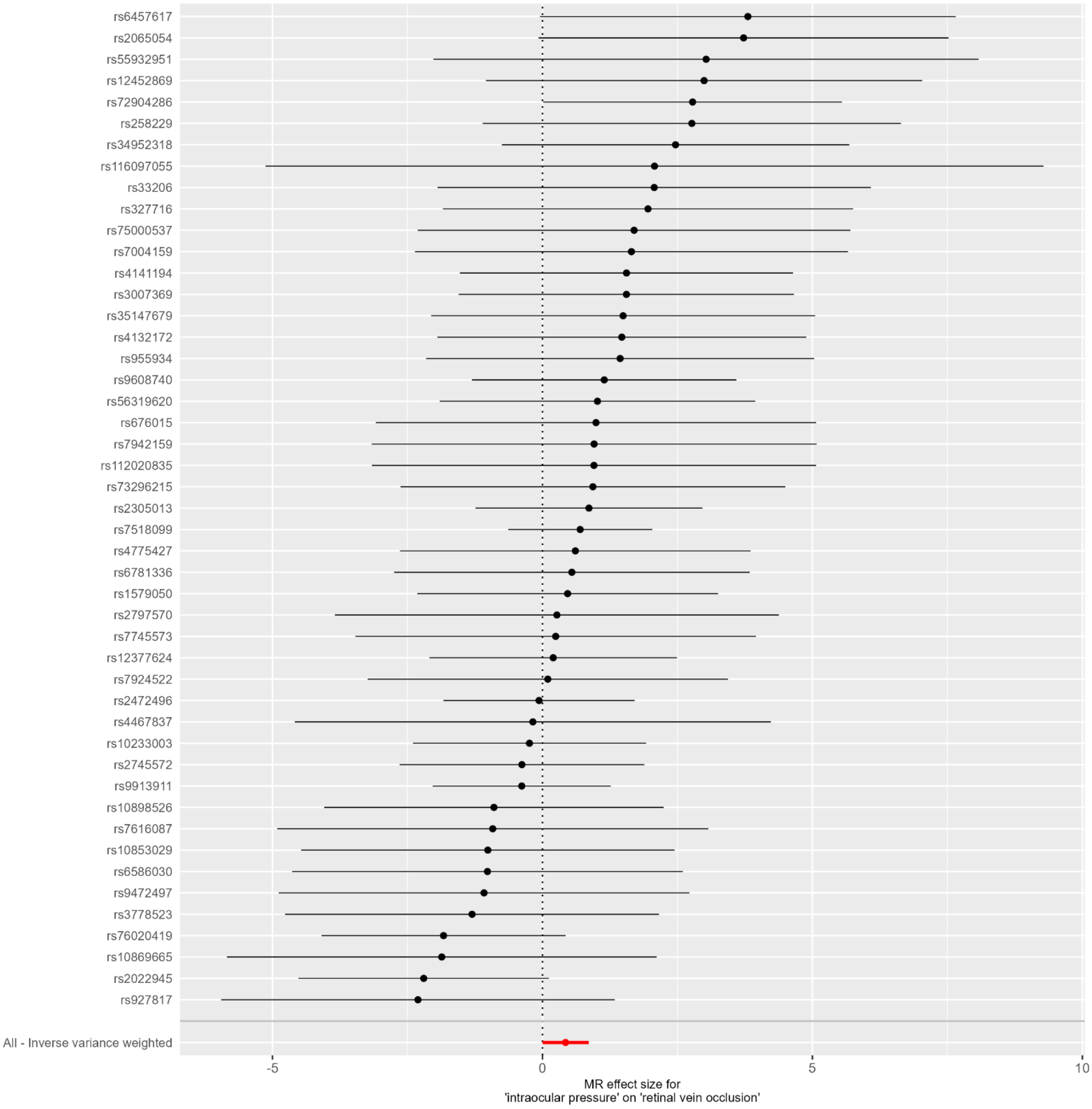


SNP: single-nucleotide polymorphism

**Supplementary Figure 2** Scatter plot of SNP-intraocular pressure associations vs SNP-retinal vein occlusion associations

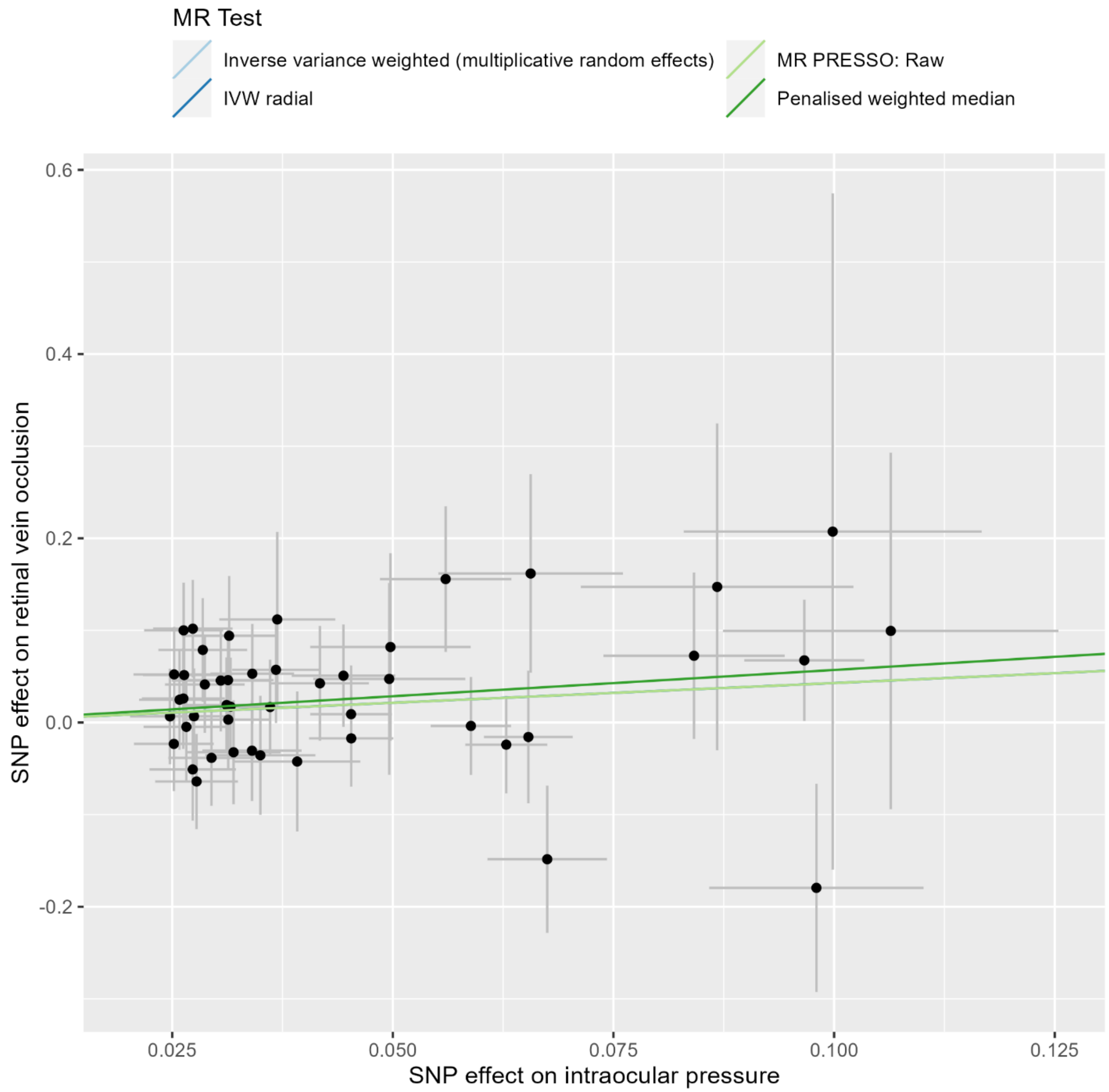


SNP: single-nucleotide polymorphism. Lines for inverse variance weighted and inverse variance weighted radial are not visible because they have the same slope as the MR PRESSO line

**Supplementary Table 1** Phenotypic descriptive statistics of studies included in the exposure, mediator, and outcome genome-wide association studies

| GWAS / Study | Phenotype | | | |
|---|---|---|---|---|
| (Gharahkhani et al., 2021) | Primary open angle glaucoma | | | |
| | N cases | N controls | Age (mean ± SD) cases | Age (mean ± SD) controls |
| NEIGHBOR/MEEI | 2606 | 2606 | 65.7 ± 13.4 | 68.3 ± 11.4 |
| EPIC-Norfolk Eye Study | 664 | 5630 | 61.5 ± 7.9 | 55.6 ± 7.9 |
| ANZRAG | 3071 | 6750 | NA | NA |
| UKBB POAG ICD10 code | 1448 | 22107 | 62.67 ± 5.6 | 56.36 ± 7.9 |
| Kaiser Permanente GERA Cohort | 3819 | 47961 | 79.6 ± 9.4 | 69.7 ± 13.0 |
| KCL | 576 | 287 | 70.5±13.5 | 80.3±4.3 |
| BMES | 107 | 600 | 68.9 ± 7.9 | 63.8 ± 8.3 |
| Southampton | 941 | 1557 | >40 | >40 |
| GHS | 47 | 2731 | 62.21 ±10.63 | 55.66 ±10.86 |
| HPFS illumina | 116 | 527 | 58.3 ± 8.4 | 51.5 ± 8.0 |
| HPFS Affy | 34 | 1297 | 58.7 ± 8.0 | 52.8 ± 7.9 |
| NHS illumina | 266 | 1692 | 55.8 ± 6.4 | 53.2 ± 6.5 |
| NHS Affy | 46 | 1992 | 55.7 ± 6.0 | 53.0 ± 6.7 |
| ERF | 110 | 1999 | NA | NA |
| Rotterdam Study I | 198 | 1282 | NA | NA |
| FINNGEN | 1824 | 93036 | 65.6 ± 11.9 | 57.8 ± 16.1 |
| Geisinger 60K | 664 | 5904 | >18 | >18 |
| Geisinger 30K | 140 | 1622 | >18 | >18 |
| | Retinal vein occlusion | | | |
| | N cases | N controls | | |
| FINNGEN (Kurki et al., 2023) | 775 | 376502 | | |
| | Intraocular pressure | | | |
| | N | | | |
| UKBB (Bycroft et al., 2018) | 97653 | | | |

**Supplementary Table 2** Associations of single nucleotide polymorphisms with intraocular pressure

| | | | Estimates for intraocular pressure | | | | | Estimates for retinal vein occlusion | | |
|---|---|---|---|---|---|---|---|---|---|---|
| SNP | EA | OA | EAF | BETA | SE | P | F | BETA | SE | P |
| Intraocular pressure | | | | | | | | | | |
| rs10233003 | A | C | 0.274 | 0.065 | 0.005 | 1.1e-38 | 169.1 | -0.016 | 0.072 | 0.827 |
| rs10853029 | T | C | 0.772 | 0.032 | 0.005 | 2.0e-09 | 35.9 | -0.032 | 0.056 | 0.566 |
| rs10869665 | T | C | 0.299 | -0.027 | 0.005 | 2.4e-08 | 31.1 | 0.051 | 0.055 | 0.358 |
| rs10898526 | A | G | 0.199 | 0.034 | 0.006 | 1.3e-09 | 36.8 | -0.031 | 0.055 | 0.575 |
| rs112020835 | A | G | 0.074 | -0.050 | 0.009 | 8.6e-09 | 33.1 | -0.047 | 0.104 | 0.649 |
| rs116097055 | T | C | 0.018 | 0.100 | 0.017 | 3.3e-09 | 35.0 | 0.207 | 0.367 | 0.572 |
| rs12377624 | C | G | 0.373 | -0.045 | 0.005 | 1.2e-22 | 95.9 | -0.009 | 0.053 | 0.865 |
| rs12452869 | T | C | 0.235 | 0.031 | 0.005 | 4.4e-09 | 34.4 | 0.094 | 0.065 | 0.146 |
| rs1579050 | G | A | 0.574 | 0.036 | 0.005 | 1.9e-15 | 63.1 | 0.017 | 0.051 | 0.743 |
| rs2022945 | G | A | 0.874 | 0.067 | 0.007 | 1.8e-23 | 99.7 | -0.148 | 0.080 | 0.063 |
| rs2065054 | T | C | 0.458 | -0.027 | 0.004 | 1.2e-09 | 36.9 | -0.102 | 0.053 | 0.055 |
| rs2305013 | T | A | 0.050 | 0.084 | 0.010 | 2.7e-16 | 67.0 | 0.072 | 0.090 | 0.422 |
| rs2472496 | A | G | 0.558 | -0.059 | 0.005 | 2.7e-38 | 167.4 | 0.004 | 0.053 | 0.944 |
| rs258229 | A | C | 0.276 | -0.028 | 0.005 | 1.5e-08 | 32.1 | -0.079 | 0.056 | 0.162 |
| rs2745572 | G | A | 0.335 | -0.045 | 0.005 | 2.3e-21 | 90.1 | 0.017 | 0.052 | 0.742 |
| rs2797570 | A | G | 0.452 | -0.025 | 0.005 | 4.2e-08 | 30.1 | -0.007 | 0.052 | 0.899 |
| rs3007369 | G | A | 0.657 | -0.034 | 0.005 | 6.2e-13 | 51.8 | -0.053 | 0.054 | 0.326 |
| rs327716 | G | A | 0.422 | -0.026 | 0.005 | 1.7e-08 | 31.9 | -0.052 | 0.051 | 0.313 |
| rs33206 | A | G | 0.605 | 0.025 | 0.005 | 4.2e-08 | 30.1 | 0.052 | 0.052 | 0.312 |
| rs34952318 | A | G | 0.049 | -0.066 | 0.010 | 3.6e-10 | 39.3 | -0.162 | 0.108 | 0.133 |
| rs35147679 | T | C | 0.204 | 0.031 | 0.006 | 4.0e-08 | 30.2 | 0.046 | 0.055 | 0.410 |
| rs3778523 | C | T | 0.319 | -0.029 | 0.005 | 1.7e-09 | 36.2 | 0.038 | 0.052 | 0.460 |

| | | | | | | | | | | |
|---|---|---|---|---|---|---|---|---|---|---|
| rs4132172 | T | C | 0.256 | -0.031 | 0.005 | 1.2e-09 | 37.0 | -0.046 | 0.055 | 0.399 |
| rs4141194 | A | C | 0.287 | 0.037 | 0.005 | 1.2e-13 | 55.0 | 0.057 | 0.058 | 0.323 |
| rs4467837 | T | C | 0.688 | -0.027 | 0.005 | 3.8e-08 | 30.3 | 0.005 | 0.060 | 0.937 |
| rs4775427 | T | C | 0.566 | 0.031 | 0.005 | 5.5e-12 | 47.5 | 0.019 | 0.052 | 0.713 |
| rs55932951 | T | C | 0.137 | 0.037 | 0.007 | 1.9e-08 | 31.6 | 0.112 | 0.095 | 0.239 |
| rs56319620 | A | C | 0.203 | 0.042 | 0.006 | 5.9e-14 | 56.4 | 0.043 | 0.062 | 0.494 |
| rs6457617 | T | C | 0.483 | 0.026 | 0.004 | 4.1e-09 | 34.6 | 0.100 | 0.052 | 0.053 |
| rs6586030 | G | A | 0.846 | 0.035 | 0.006 | 2.0e-08 | 31.5 | -0.036 | 0.065 | 0.581 |
| rs676015 | C | T | 0.630 | -0.026 | 0.005 | 2.2e-08 | 31.4 | -0.026 | 0.055 | 0.634 |
| rs6781336 | G | A | 0.298 | -0.032 | 0.005 | 1.3e-10 | 41.3 | -0.017 | 0.053 | 0.746 |
| rs7004159 | T | C | 0.065 | 0.050 | 0.009 | 4.4e-08 | 30.0 | 0.082 | 0.102 | 0.421 |
| rs72904286 | T | C | 0.102 | -0.056 | 0.007 | 5.4e-14 | 56.6 | -0.156 | 0.079 | 0.049 |
| rs73296215 | C | T | 0.014 | -0.106 | 0.019 | 2.1e-08 | 31.4 | -0.099 | 0.194 | 0.607 |
| rs75000537 | T | C | 0.021 | -0.087 | 0.015 | 1.9e-08 | 31.6 | -0.147 | 0.177 | 0.406 |
| rs7518099 | T | C | 0.876 | -0.097 | 0.007 | 6.9e-46 | 202.2 | -0.067 | 0.066 | 0.305 |
| rs76020419 | T | G | 0.035 | -0.098 | 0.012 | 6.8e-16 | 65.2 | 0.180 | 0.113 | 0.112 |
| rs7616087 | T | C | 0.441 | 0.025 | 0.005 | 2.4e-08 | 31.1 | -0.023 | 0.051 | 0.651 |
| rs7745573 | C | A | 0.444 | -0.027 | 0.005 | 1.2e-09 | 37.0 | -0.007 | 0.052 | 0.897 |
| rs7924522 | A | C | 0.662 | 0.031 | 0.005 | 3.9e-11 | 43.7 | 0.003 | 0.053 | 0.954 |
| rs7942159 | G | A | 0.587 | -0.026 | 0.005 | 1.6e-08 | 31.9 | -0.025 | 0.054 | 0.649 |
| rs927817 | C | T | 0.361 | 0.028 | 0.005 | 3.1e-09 | 35.1 | -0.064 | 0.052 | 0.215 |
| rs9472497 | G | A | 0.109 | 0.039 | 0.007 | 4.1e-08 | 30.1 | -0.042 | 0.076 | 0.577 |
| rs955934 | A | C | 0.456 | 0.029 | 0.005 | 1.9e-10 | 40.6 | 0.041 | 0.053 | 0.433 |
| rs9608740 | C | A | 0.190 | 0.044 | 0.006 | 3.5e-14 | 57.4 | 0.051 | 0.056 | 0.360 |
| rs9913911 | G | A | 0.374 | -0.063 | 0.005 | 8.8e-42 | 183.4 | 0.024 | 0.053 | 0.649 |

EA, effect allele. OA, other allele. EAF, effect allele frequency. SE, standard error. SNPs rs13024279 and rs2386136 were present only in the UKBB GWAS.

**Supplementary Table 3** Association ($P<5x10^{-8}$) of the single nucleotide polymorphisms used as instruments with confounders or outcome risk factors in PhenoScanner (accessed on 2023/11/20 using the phenoscanner function of the R MendelianRandomization package)

| SNP | Phenotypes | PMID |
|---|---|---|
| Intraocular pressure | | |
| rs2472496 | Glaucoma primary open angle | rs2472496 |
| rs56319620 | Lymphocyte count | rs56319620 |
| rs56319620 | High density lipoprotein | rs56319620 |
| rs6457617 | Eosinophil count | rs6457617 |
| rs6457617 | Eosinophil percentage of granulocytes | rs6457617 |
| rs6457617 | Eosinophil percentage of white cells | rs6457617 |
| rs6457617 | Granulocyte count | rs6457617 |
| rs6457617 | Lymphocyte count | rs6457617 |
| rs6457617 | Mean platelet volume | rs6457617 |
| rs6457617 | Monocyte count | rs6457617 |
| rs6457617 | Myeloid white cell count | rs6457617 |
| rs6457617 | Neutrophil count | rs6457617 |
| rs6457617 | Sum basophil neutrophil counts | rs6457617 |
| rs6457617 | Sum eosinophil basophil counts | rs6457617 |
| rs6457617 | Sum neutrophil eosinophil counts | rs6457617 |
| rs6457617 | White blood cell count | rs6457617 |
| rs6457617 | IgA deficiency | rs6457617 |
| rs6457617 | Height | rs6457617 |

| | | |
|---|---|---|
| rs6457617 | Alopecia areata | rs6457617 |
| rs6457617 | Idiopathic membranous nephropathy | rs6457617 |
| rs6457617 | Multiple sclerosis | rs6457617 |
| rs6457617 | Rheumatoid arthritis | rs6457617 |
| rs6457617 | Rheumatoid arthritis ACPA positive | rs6457617 |
| rs6457617 | Rheumatoid arthritis combined control dataset | rs6457617 |
| rs6457617 | Systemic sclerosis | rs6457617 |
| rs6457617 | Systemic sclerosis Anti centromere antibody negative | rs6457617 |
| rs6457617 | Systemic sclerosis Anti centromere antibody positive | rs6457617 |
| rs6457617 | Systemic sclerosis Anti topoisomerase I antibody negative | rs6457617 |
| rs6457617 | Systemic sclerosis Anti topoisomerase I antibody positive | rs6457617 |
| rs6457617 | Systemic sclerosis Diffuse cutaneous cases | rs6457617 |
| rs6457617 | Systemic sclerosis Limited cutaneous cases | rs6457617 |
| rs6457617 | Systemic lupus erythematosus | rs6457617 |
| rs6457617 | Primary sclerosing cholangitis | rs6457617 |
| rs6457617 | Immunoglobulin G index levels in multiple sclerosis | rs6457617 |
| rs6457617 | Arthritis rheumatoid | rs6457617 |
| rs6457617 | Lupus erythematosus systemic | rs6457617 |
| rs6457617 | Scleroderma systemic | rs6457617 |
| rs6457617 | Sclerosis | rs6457617 |
| rs6586030 | Inflammatory bowel disease | rs6586030 |

| | | | |
|---|---|---|---|
| rs7518099 | Open angle glaucoma | | rs7518099 |

PMID, PubMed ID.

**Supplementary Table 4** Heterogeneity of Wald ratios and MR-Egger test for directional pleiotropy

| Intraocular pressure | Heterogeneity | | | |
|---|---|---|---|---|
| | Q | Degrees of Freedom | P | $I_{GX}^2$ |
| Intraocular pressure | 37.89 | 46 | 0.797 | 0 |
| | MR-Egger test for directional pleiotropy | | | |
| | Intercept | Standard error | P | |
| Intraocular pressure | 0.025 | 0.023 | 0.27 | |

**Supplementary Table 5** Inverse variance weighted estimates in leave-one-out analysis in primary analysis

| SNP excluded | SNP | OR | (95% CI) | P value |
|---|---|---|---|---|
| Intraocular pressure | rs10233003 | 1.58 | (1.06;2.35) | 0.025 |
| | rs10853029 | 1.57 | (1.06;2.32) | 0.024 |
| | rs10869665 | 1.58 | (1.07;2.32) | 0.022 |
| | rs10898526 | 1.57 | (1.06;2.33) | 0.024 |
| | rs112020835 | 1.53 | (1.03;2.26) | 0.036 |
| | rs116097055 | 1.53 | (1.03;2.26) | 0.035 |
| | rs12377624 | 1.55 | (1.04;2.31) | 0.032 |
| | rs12452869 | 1.49 | (1.01;2.19) | 0.043 |
| | rs1579050 | 1.53 | (1.03;2.28) | 0.035 |
| | rs2022945 | 1.68 | (1.16;2.44) | 0.006 |
| | rs2065054 | 1.47 | (1.01;2.15) | 0.046 |
| | rs2305013 | 1.51 | (1.01;2.25) | 0.045 |
| | rs2472496 | 1.58 | (1.06;2.37) | 0.026 |
| | rs258229 | 1.49 | (1.01;2.2) | 0.043 |
| | rs2745572 | 1.58 | (1.06;2.35) | 0.024 |
| | rs2797570 | 1.54 | (1.04;2.28) | 0.033 |
| | rs3007369 | 1.50 | (1.01;2.23) | 0.043 |
| | rs327716 | 1.51 | (1.02;2.23) | 0.041 |
| | rs33206 | 1.51 | (1.02;2.23) | 0.040 |
| | rs34952318 | 1.48 | (1;2.18) | 0.048 |
| | rs35147679 | 1.51 | (1.02;2.24) | 0.040 |
| | rs3778523 | 1.58 | (1.07;2.33) | 0.022 |

| | | | |
|---|---|---|---|
| rs4132172 | 1.51 | (1.02;2.24) | 0.040 |
| rs4141194 | 1.50 | (1.01;2.23) | 0.043 |
| rs4467837 | 1.54 | (1.04;2.29) | 0.031 |
| rs4775427 | 1.53 | (1.03;2.27) | 0.035 |
| rs55932951 | 1.51 | (1.02;2.22) | 0.039 |
| rs56319620 | 1.52 | (1.02;2.25) | 0.040 |
| rs6457617 | 1.47 | (1.01;2.15) | 0.046 |
| rs6586030 | 1.57 | (1.06;2.32) | 0.025 |
| rs676015 | 1.53 | (1.03;2.26) | 0.036 |
| rs6781336 | 1.53 | (1.03;2.28) | 0.035 |
| rs7004159 | 1.51 | (1.02;2.24) | 0.039 |
| rs72904286 | 1.45 | (0.99;2.12) | 0.057 |
| rs73296215 | 1.52 | (1.03;2.26) | 0.037 |
| rs75000537 | 1.51 | (1.02;2.24) | 0.039 |
| rs7518099 | 1.49 | (0.98;2.25) | 0.060 |
| rs76020419 | 1.67 | (1.14;2.44) | 0.008 |
| rs7616087 | 1.56 | (1.05;2.31) | 0.027 |
| rs7745573 | 1.54 | (1.04;2.29) | 0.033 |
| rs7924522 | 1.54 | (1.04;2.29) | 0.032 |
| rs7942159 | 1.53 | (1.03;2.26) | 0.036 |
| rs927817 | 1.59 | (1.09;2.34) | 0.017 |
| rs9472497 | 1.57 | (1.06;2.32) | 0.025 |
| rs955934 | 1.51 | (1.02;2.24) | 0.039 |
| rs9608740 | 1.50 | (1.01;2.23) | 0.045 |

| | | | | |
|---|---|---|---|---|
| | rs9913911 | 1.63 | (1.09;2.43) | 0.017 |
| | All | 1.53 | (1.04;2.26) | 0.031 |

**Supplementary Table 6** Mediation effect of intraocualr pressure on retinal vein occlusion via primary open-angle glaucoma.

| Exposure | Mediator | Outcome | Total effect of IOP on RVO | Effect of exposure (IOP) on mediator (POAG) | Effect of mediator (POAG) on outcome (RVO) | Mediation effect of IOP on RVO via POAG | | Mediated proportion |
|---|---|---|---|---|---|---|---|---|
| | | | Effect size (95% CI) | Effect size (95% CI) | Effect size (95% CI) | Effect size (95% CI) | P value | % |
| Intraocular pressure | Primary open-angle glaucoma | Retinal vein occlusion | 0.429 (0.040 to 0.817) | 1.644 (1.227 to 2.061) | 0.239 (0.071 to 0.407) | 0.393 (0.099 to 0.686) | 0.009 | 91.6 |

RVO: retinal vein occlusion; POAG: primary open-angle glaucoma; IOP: intraocular pressure; All estimates are shown as the difference in the logarithm of odds of the outcome, per 1 unit increase of the exposure (continuous variables: IOP) or between the two exposure groups (binary variables: POAG)
